# A Wearable Platform for Real-Time Control of a Prosthetic Hand by High-Density EMG

**DOI:** 10.1101/2025.10.08.25337421

**Authors:** Ricardo G. Molinari, Valeria Avilés-Carrillo, Guilherme A. G. De Villa, Leonardo A. Elias

## Abstract

This study presents a heterogeneous embedded architecture that addresses a fundamental gap in wearable myoelectric systems: the inability of existing platforms to simultaneously provide high-density signal acquisition, computational flexibility, and autonomy. The platform integrates two 64-channel RHD2164 front-ends (128 channels total) with a Zynq UltraScale+ multiprocessor system-on-chip for heterogeneous processing. A PYNQ-based Python/Linux framework enables scalable algorithm development. Experiments with 21 healthy subjects performing eight motor tasks (finger flexion/extension, thumb opposition, and grasp patterns) at two frequencies (0.50 and 0.75 Hz) demonstrated the platform’s capability in high-density surface electromyography (HD sEMG) recording and real-time control of a single degree of freedom (1-DoF). Signal quality exceeded recommended thresholds (Signal-to-Noise Ratio: 13.93 ± 7.51 dB; Signal-to-Motion-artifact Ratio: 25.18 ± 5.18 dB), confirming the effectiveness of the dual-front-end architecture. The processing pipeline combined reinforced electrode signal adaptation (RESA), non-negative matrix factorization (NMF), and Kalman filtering, resulting in strong agreement between estimated and reference signals, with maximum normalized cross-correlation (XC_max_) values from 0.54 ± 0.22 to 0.80 ± 0.16. The coefficient of determination (R^2^) for HD sEMG reconstruction ranged from 0.87 ± 0.09 to 0.95 ± 0.03, with higher values for prehension tasks. End-to-end latency from acquisition to command output ranged from 63.3 ± 1.0 ms (30 ms buffer) to 219.1 ± 4.5 ms (150 ms buffer), maintaining temporal alignment (XC_max_ lag: 0.02 ± 2.09 s). The heterogeneous architecture supports full local processing, with the FPGA handling acquisition and the Arm Cortex-A53 cores performing motor intention decoding, providing a scalable foundation for adaptive multi-DoF prosthetic control.

## I. INTRODUCTION

HIGH-DENSITY surface electromyography (HD sEMG) has become a key technology for non-invasive neuro-muscular signal acquisition, enabling advanced control of upper-limb prostheses [1]. By capturing spatially detailed muscle activation patterns with dense electrode grids, HD sEMG allows a more accurate decoding of user intent, which is essential for intuitive and precise prosthetic control. This is particularly relevant for transradial amputees, for whom restoring fine finger movements and proportional grip modulation remains a major challenge [2]. Nevertheless, the rejection rate of below-elbow prostheses remains as high as 71.4 %, mainly owing to issues of comfort, weight, and limited functionality [3].

The technological evolution of myoelectric control systems has followed a clear trajectory toward increasingly embedded and autonomous processing architectures. Early real-time control schemes relied on desktop computers connected to external amplifiers, offering limited portability and scalability [4]. The advent of wireless communication enabled cloud-based architectures, in which wearable acquisition devices transmitted raw signals to remote servers for computation [5]. However, this paradigm introduced critical limitations, including dependency on network reliability, increased end-to-end latency, elevated power consumption, and privacy concerns related to the transmission of physiological data [6].

Hybrid and distributed architectures subsequently emerged as an intermediate solution, in which part of the signal processing is performed locally on the wearable device while more computationally demanding tasks are offloaded to external devices such as smartphones, gateways, or nearby servers [7], [8]. Although this approach reduces communication bandwidth compared to fully cloud-based systems, it still depends on external computing resources and, therefore, on continuous connectivity, which prevents fully autonomous operation and precludes deterministic real-time behavior in the absence of a reliable communication link.

The current frontier lies in fully embedded processing, where high-density signal acquisition and advanced decoding algorithms are executed entirely on the device. In this configuration, the wearable platform itself becomes the true computational ‘‘edge’’, enabling autonomous operation, minimal end-to-end latency, predictable timing behavior, and reliable closed-loop control without dependence on external hardware or network connectivity [9]–[11].

However, achieving fully embedded HD sEMG processing introduces a fundamental hardware architectural challenge that existing platforms have been unable to resolve. Current wearable systems fall into one of two categories, each with critical structural limitations. Microcontroller (MCU)-centered platforms provide low power consumption, compact form factors, and ease of programming, but their sequential processing architecture and limited I/O bandwidth and memory throughput make them fundamentally unsuitable for scaling to high-density electrode arrays while maintaining deterministic real-time behavior under concurrent processing loads [12], [13]. Conversely, field-programmable gate array (FPGA)-centered platforms excel at deterministic data acquisition and parallel processing, but their hardware-centric programming model severely restricts algorithmic flexibility, increases development complexity, and provides poor support for high-level tasks such as system orchestration, user interfaces, and integration of modern machine learning frameworks [13], [14].

The fundamental hardware architectural gap can be stated as follows: existing wearable platforms cannot simultaneously provide (i) deterministic, scalable acquisition of large-scale electrode arrays (100+ channels), (ii) sufficient computational flexibility to support rapid development and deployment of advanced decoding algorithms, including deep learning models, and (iii) autonomy within the strict power, size, and thermal constraints of wearable systems. MCU-based architectures are constrained by inherent bandwidth and processing limitations, preventing compliance with requirements (i) and (ii); FPGA-only architectures, while offering high throughput, fall short of requirement (ii) due to limited software programmability and lack of high-level abstraction; and hybrid or distributed architectures that offload computation to external devices compromise requirement (iii) by sacrificing autonomy and introducing unpredictable latencies. This architectural mismatch represents a critical barrier to translating state-of-the-art HD sEMG decoding algorithms from laboratory environments to real-world wearable prosthetic systems.

This architectural gap becomes critical when confronted with the computational requirements of state-of-the-art HD sEMG algorithms. Recent advances in HD sEMG processing have demonstrated the feasibility of non-invasive motor unit decomposition and the importance of robustness to non-stationary signal conditions [15]. Modern decoding strategies based on deep learning have achieved remarkable performance in proportional and simultaneous control of multiple degrees of freedom (DoF) [16]. However, both approaches impose substantial computational demands, including large-scale matrix operations, iterative optimization procedures, and significant memory requirements. Without a hardware architecture that can reconcile deterministic acquisition with computational flexibility, these algorithms remain confined to desktop environments where resources are virtually unconstrained, preventing their deployment in practical wearable prostheses [11], [17]. Thus, the challenge is not merely algorithmic, but fundamentally architectural: bridging the gap between what algorithms can achieve in controlled laboratory settings and what embedded hardware can execute in realworld wearable applications.

This work directly addresses this architectural mismatch by presenting a 128-channel HD sEMG acquisition and processing platform based on the Zynq UltraScale+ multiprocessor system-on-chip (MPSoC), which serves simultaneously as a functional wearable system and as a reference architectural model for next-generation myoelectric control. While Zynq-based platforms have been previously explored for myoelectric applications, to the best of our knowledge, this is the first to integrate the high-performance UltraScale+ architecture, featuring programmable logic tightly coupled with a quad-core Arm Cortex-A53 APU and additional hardware accelerators, with the PYNQ framework. This combination uniquely enables deterministic FPGA-based acquisition, sufficient computational power for advanced decoding algorithms, and rapid prototyping capabilities typically restricted to desktop environments, enabling experimental evaluation of state-of-the-art methods under realistic wearable constraints.

## II. ACQUISITION AND PROCESSING SYSTEM

The HD sEMG acquisition and processing platform was conceived as a heterogeneous embedded architecture that explicitly separates deterministic time-critical signal acquisition from flexible high-level processing tasks. This design choice addresses two fundamental requirements of high-density myoelectric systems: (i) reliable and low-latency acquisition of a large number of channels, and (ii) sufficient computational and software flexibility to support real-time decoding, user interaction, and system-level control. By combining dedicated hardware acquisition with a programmable processing environment, the proposed architecture overcomes the limitations of MCU-only platforms, which typically face scalability and real-time performance constraints at high channel counts, while also avoiding the higher development effort and longer iteration cycles typically required by purely FPGA-based solutions when implementing complex signal processing pipelines, adaptive decoding algorithms, and user-facing interfaces, despite their intrinsic flexibility and performance [18].

Fig. 1 illustrates the overall system architecture. The platform consists of two analog front-ends and a heterogeneous computing system implemented on a Zynq UltraScale+ Multiprocessor System-on-Chip (MPSoC, Advanced Micro Devices Inc., USA). The analog front-ends are based on RHD2164 devices (Intan Technologies, USA), each connected to a 64-channel electrode grid (GR10MM0808, OT Bioelettronica, Italy). These integrated front-ends were selected because they provide low-noise amplification, programmable filtering, high-resolution digitization, and synchronized multichannel data streaming in a compact form factor, which is particularly suitable for scalable HD sEMG acquisition. Using two independent 64-channel front-ends enables straightforward expansion to 128 channels while preserving signal integrity and acquisition determinism.

**FIGURE 1.**
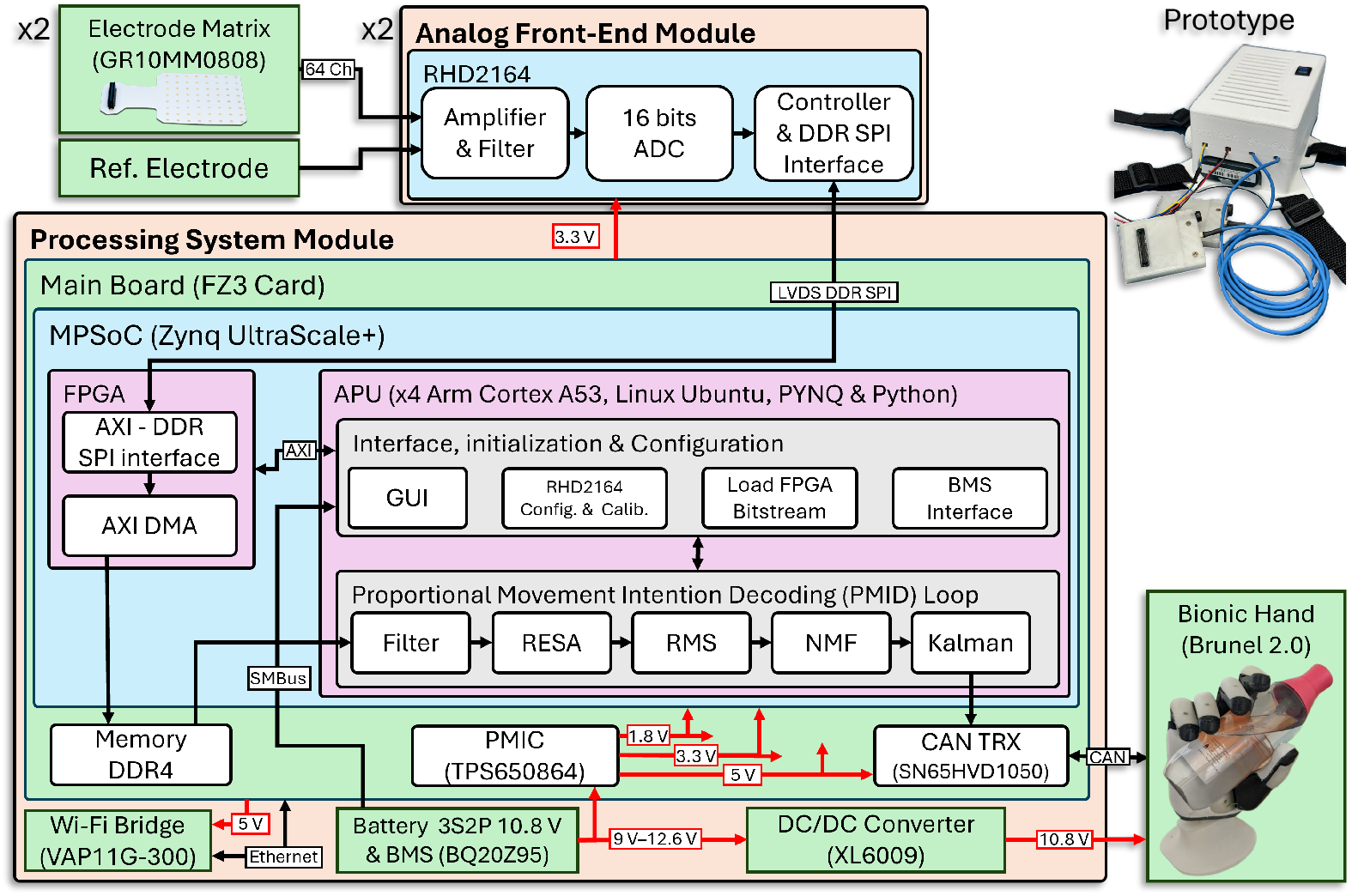
Schematic diagram of the developed HD sEMG acquisition and processing platform for real-time prosthetic hand control. Two 64-channel analog front-ends amplify, filter, digitize, and transmit the signals to a heterogeneous processing system (Zynq UltraScale+ MPSoC), which combines FPGA and CPU cores running a customized Linux-based PYNQ framework. The system handles signal acquisition, movement intention decoding, graphical user interface (GUI) operation, and battery monitoring. A rechargeable Li-ion battery pack with a battery management system (BMS) powers the platform and the prosthetic hand, with voltage regulated for each device. The image illustrates the hardware and software components as well as the information and power flow. Wireless access to the GUI is provided via a Wi-Fi bridge. The top-right corner shows the platform prototype, and the bottom-right corner shows the Brunel Hand 2.0 (Open Bionics, UK) controlled by the platform.

The processing system is implemented on the FZ3 CARD development board (MYiR Tech Ltd., China), which integrates a Zynq UltraScale+ MPSoC. The choice of an MPSoC, rather than a conventional high-performance MCU or a discrete FPGA–MCU combination, was motivated by several architectural advantages. In particular, the MPSoC architecture naturally supports a clean partitioning of hard real-time acquisition tasks, real-time control functions, and computationally intensive but less time-critical processing, which is central to the proposed system and enables future extensions to more complex, computationally demanding algorithms.

First, the tight integration of the FPGA and the application processing unit (APU) within a single chip provides a high-bandwidth, low-latency communication infrastructure through shared memory and direct memory access (DMA), eliminating the overhead and power consumption associated with external interfaces commonly used in discrete solutions. Beyond the APU–FPGA pair, the MPSoC also integrates a Mali-400 graphics processing unit (GPU) and dual Arm Cortex-R5F real-time cores, which further extend the computational heterogeneity of the platform and enable the offloading of computationally demanding tasks, as well as the implementation of future control and signal processing pipelines of increased complexity. Second, this integration results in a more compact and energy-efficient solution, which is essential for wearable and portable applications. Third, it simplifies hardware design and improves system reliability by reducing the number of discrete components and potential points of failure during manufacturing.

Within this architecture, the FPGA is responsible for all time-critical acquisition and data transfer. It acts as a protocol bridge between the analog front-ends and the processing system, managing the synchronous reception of digitized sEMG data streams from both RHD2164 devices. The FPGA handles data framing, buffering, and DMA transfers to shared memory, thereby ensuring deterministic timing behavior and minimizing acquisition latency and jitter. This hardware-based acquisition pipeline is essential for HD sEMG systems, where a large number of channels and high sampling rates make purely software-based acquisition impractical. Moreover, implementing acquisition in the FPGA allows the platform to scale to higher channel numbers without increasing CPU load, which is a key limitation of MCU-centered approaches reported in the literature [12], [13]. In the proposed platform, this functionality is materialized through a dedicated hardware interface that implements the complete communication and data transfer pipeline between the RHD2164 devices and the MPSoC memory subsystem.

### A. THE ANALOG FRONT-END MODULE

The analog front-end (AFE) module represents the first stage of the deterministic acquisition pipeline and is responsible for conditioning and digitizing the raw biopotential signals, ensuring signal integrity, low-noise amplification, and precise digitization before their transfer to the FPGA-based hardware interface. Tab. 1 presents the critical parameters for HD sEMG acquisition and compares the recommended features [19], [20] with the technical specifications of the RHD2164 device (as provided in its datasheet). The comparison includes aspects of analog front-end quality (such as amplitude, impedance, noise rejection, voltage range, intrinsic noise, and filtering) and analog-to-digital converter (ADC) performance, including the sampling rate and resolution.

The device employed in the platform features configurable third-order Butterworth analog band-pass filters, whose cut-off frequencies were set to 10 Hz and 500 Hz [21]. Moreover, a first-order digital high-pass filter was activated and configured with a cutoff frequency of 10 Hz to remove low-frequency oscillations and baseline drift. The RHD2164 operates in slave mode with a sampling rate of 2052.52 Hz per channel, which is directly determined by the frequency of the conversion commands generated by the FPGA via the Serial Peripheral Interface (SPI). Each RHD2164 integrates two 32-channel modules that operate in parallel, with the ADC data combined using a Double Data Rate (DDR) multiplexer for transmission via the Master Input Slave Output (MISO) channel. Low-Voltage Differential Signaling (LVDS) was employed for data transmission through 0.9-meter flexible cables (RHD SPI Interface, Intan Technologies, USA) connecting the analog front-ends to the processing system.

### B. DUAL-MODE FPGA INTERFACE FOR RHD2164 DEVICES

1. This module constitutes the core of the deterministic acquisition pipeline of the proposed platform, implementing in hardware the time-critical communication between the RHD2164 analog front-ends and the shared memory sub-system of the MPSoC. The Advanced Extensible Interface (AXI)–DDR SPI module, implemented in the MPSoC FPGA, converts between the LVDS DDR SPI protocol used by the RHD2164 devices and the AXI protocol used internally by the processing system. A finite-state machine is employed to manage the bridge between these protocols, handle data alignment across the master DDR SPI interface channels, perform (de)serialization, identify valid MISO message cycles, and control the interface’s operating modes.

**TABLE 1.** Comparison between recommended HD sEMG signal acquisition features and RHD2164 specifications.

| Specification | Recommendations [19], [20] | RHD2164 |
| --- | --- | --- |
| Amplifier AC Input Voltage Range, mV | $\pm 7.5$ | $\pm 5.0$ |
| Input Impedance, MΩ | $\geq 80$ at 50–60 Hz | 1300 at 10 Hz and 13 at 1 kHz |
| Common-Mode Rejection Ratio (CMRR), dB | $\geq 90$ at 50–60 Hz | 82 at 50–60 Hz |
| Power Supply Rejection Ratio (PSRR), dB | $\geq 60$ | 75 |
| Input-Referred Noise, $\mu V_{RMS}$ | $\leq 2.0$ | 2.4 |
| High-Pass Cutoff Frequency (analog prefilter), Hz | 0.10–20 | 0.02–1000 (set to 10) |
| Low-Pass Cutoff Frequency (analog prefilter), Hz | 500–1000 | 10–20000 (set to 500) |
| ADC Sampling Rate per Channel, Hz | $\geq 2048$ | up to 30000 (set to 2052.52) |
| ADC Resolution, bits | $\geq 12$ | 16 |
| Input-Referred Resolution, $\mu V$ | $\leq 0.500$ | 0.195 |

Two operating modes are supported: configuration and streaming. In configuration mode, the interface enables the graphical user interface (GUI), running on the APU, to access the internal registers of the RHD2164 devices for setup, calibration, and parameter tuning, thereby decoupling device configuration from the real-time acquisition pipeline. In streaming mode, the module continuously generates analog- to-digital conversion commands for the RHD2164 devices and transfers the acquired multichannel data to external memory through the AXI DMA block, ensuring uninterrupted and deterministic data flow independently of the processing load imposed on the APU.

The AXI DMA operates cyclically based on the buffer size defined through the APU GUI, allowing flexible adjustment of memory allocation while sustaining a continuous high-throughput data stream. This hardware-based data transfer mechanism is designed to provide predictable latency and efficient memory subsystem utilization, both essential requirements for real-time HD sEMG acquisition. The FPGA interface was designed to comply with the timing constraints of the RHD2164 communication protocol, with setup and hold times of 10.40 ns and 12 ns, respectively, both compliant with the timing requirements specified in the device datasheet. These design characteristics ensure reliable communication between the analog front-ends and the MPSoC, supporting the FPGA’s role as the backbone of the deterministic acquisition pipeline.

### C. APPLICATION PROCESSING UNIT: HIGH-LEVEL PROCESSING AND SYSTEM CONTROL

The APU, which consists of four Arm Cortex-A53 cores, executes higher-level processing and system orchestration tasks under a Linux Ubuntu operating system based on a customized PYNQ (Python Productivity for Zynq) framework. This environment was selected to provide a flexible, developer-friendly platform for implementing signal processing algorithms, movement-intention decoding, system configuration, and user interaction. In contrast to FPGA-only solutions, which often require long development cycles and specialized hardware description languages, the APU enables rapid prototyping, straightforward algorithm modification, and seamless integration of visualization and control interfaces. This combination of hardware-level determinism from the FPGA and software-level flexibility from the APU constitutes a central advantage of the proposed architecture.

Specifically, the APU is responsible for configuring the analog front-ends, initializing and controlling the FPGA logic, executing the proportional movement intention decoding (PMID) loop, hosting the GUI, and monitoring system-level parameters such as battery status. By offloading acquisition and data transfer to the FPGA, the APU can dedicate its computational resources to signal processing and control tasks without being burdened by strict real-time I/O constraints. This clear functional partitioning enhances fault containment and reduces timing interference between acquisition and processing tasks, ensuring bounded latency and deadline compliance even under full-load conditions.

The estimated proportional movement intention is transmitted to the prosthetic hand via an SN65HVD1050 transceiver (Texas Instruments, USA) using the Controller Area Network (CAN) protocol. CAN was selected for its robust error detection strategy, low latency, and widespread adoption in embedded control systems, making it well-suited for reliable communication between the decoding platform and the actuation system [22]. The target device is a modified version of the open-source Brunel Hand 2.0 (Open Bionics, UK), which enables direct integration and experimental validation of real-time prosthetic control.

The system’s GUI was developed in Jupyter Notebook and is accessed remotely through a VAP11G-300 Wi-Fi bridge (Vonets Network Co. Ltd., China). This interface enables device configuration, platform operational control, and real-time visualization of HD sEMG signals. The use of a browser-based GUI further enhances portability and usability, enabling the platform to be operated from any device on the same network without additional software installation.

### D. POWER SUPPLY AND ENERGY MANAGEMENT

The platform is powered by rechargeable lithium-ion batteries (model 369108, Hamilton Medical, Switzerland) configured as a 3S2P pack, providing a nominal voltage of 10.8 V and a total energy capacity of 72 Wh. The BQ20Z95 (Texas Instruments, USA) battery management system (BMS) ensures safe operation, charging control, and protection. The FZ3 CARD includes a power management integrated circuit (TPS650864, Texas Instruments, USA), which regulates and distributes power to the MPSoC and onboard peripherals. In parallel, the same battery pack independently powers the prosthetic hand via a Buck-Boost DC/DC converter (XL6009, KylinChip Electronic, China), which regulates the output voltage to 10.8 V.

This power architecture was designed to support continuous operation under full acquisition and processing load while preserving functional and electrical decoupling between the sensing/processing platform and the actuation system. It also provides the electrical foundation for the quantitative characterization of power consumption and runtime presented in the experimental evaluation.

### E. THE PROPORTIONAL MOVEMENT INTENTION DECODING LOOP

The PMID loop utilizes successive and overlapping analysis windows of HD sEMG recordings with a duration of *T*_*a*_ to maximize the processing efficiency of the APU [4]. The first stage of the loop implements a fourth-order Butterworth digital band-pass filter (10–500 Hz) and a 60 Hz notch filter to suppress motion artifacts, high-frequency noise, and power line interference that may not be fully attenuated by the analog front-end filtering stage. This redundant filtering strategy enhances signal robustness to interferences under real acquisition conditions.

The second stage applies the Reinforced Electrode Signal Adaptation (RESA) algorithm to mitigate non-stationary artifacts caused by impedance variations at the skin–electrode interface [23]. The RESA algorithm computes an adaptation weight matrix based on the similarity between a virtual reference signal (VRS) and the individual electrode signals. To enable real-time execution on the proposed embedded platform, two modifications were introduced to the original formulation [23] to reduce the computational complexity. First, the Beta distribution used in the VRS calculation was replaced with a Gaussian distribution. Second, the mutual information metric was substituted by Pearson’s correlation coefficient. These adaptations preserve the core functionality of the RESA algorithm while significantly improving its suitability for real-time implementation.

In the third stage, the root mean square (RMS) value of each HD sEMG channel is computed using sliding windows of duration *T*_*a*_, providing an estimation of muscle activation amplitude. The fourth stage decomposes the resulting feature matrix into two components using non-negative matrix factorization (NMF) [24]. Although NMF is a linear decomposition technique, its use is well established in myoelectric control due to its computational efficiency, numerical stability, and its ability to provide an interpretable low-dimensional representation of muscle activation patterns (muscle synergies) [25], which is particularly advantageous for real-time embedded systems. One component of the NMF decomposition captures activation patterns associated with finger flexion, thumb opposition, or grasp-closing movements, whereas the other represents finger extension, thumb reposition, or grasp-opening movements. The algorithm assigns each component to a movement direction by analyzing the phase of the cross-spectral density between its temporal activation signal and the task command signal (described in Section III-B). This directional mapping subsequently derives the proportional movement intention as the difference between the two components and standardizes the resulting signal to have zero mean and unit variance. It is worth noting that this formulation assumes a quasi-linear relationship between muscle activation synergies and movement intention, which represents a deliberate trade-off between physiological modeling fidelity and the computational feasibility required for real-time operation.

The fifth and final stage applies a one-dimensional Kalman filter [26], without an explicit dynamic model, to reduce residual noise in the estimated movement intention. The process noise variance was set to 10^−4^ and the measurement noise variance to 10^−2^, providing an effective balance between smoothing and responsiveness. This final filtering step smooths the proportional control signal while preserving the temporal resolution required for intuitive, responsive prosthetic actuation.

## III. EXPERIMENTAL EVALUATION

### A. PARTICIPANTS

Twenty-one healthy participants (10 women and 11 men) were included in the study. Their demographics (mean ± standard deviation) were: 27.19 ± 6.55 years, body weight of 73.52 ± 16.06 kg, height of 1.68 ± 0.11 m, body mass index (BMI) of 25.73 ± 3.57 kg/m^2^, hand length of 18.36 ± 1.98 cm, and forearm circumference of 26.67 ± 2.72 cm.

The BMI was computed as body weight (kg) divided by height squared (m^2^). Hand length was measured as the linear distance from the midstylion to the dactylion of the third digit using a flexible, non-elastic measuring tape with 0.1 cm resolution, following standard anthropometric procedures [27]. Forearm circumference was measured at the point of maximum girth of the dominant forearm, with the elbow extended, forearm supinated, and muscles relaxed, using the same non-elastic measuring tape applied without tissue compression [27].

Only one participant reported being left-handed. None of the participants had a history of neuromuscular or joint disorders. This study was approved by the Research Ethics Committee of the University of Campinas (CAAE 80299124.2.1001.5404) on October 24, 2024, and all participants provided informed consent before the experiments.

### B. EXPERIMENTAL PROTOCOL

The two 64-electrode grids (8×8 configuration) were placed centrally over the Extensor Digitorum Communis (EDC) and Flexor Digitorum Superficialis (FDS) muscles, following a standardized protocol [28]. The participant was seated with the forearm resting on a table throughout the electrode placement procedure. Before electrode placement, skin preparation was performed through cutaneous abrasion with an exfoliating paste followed by cleansing with 70 % alcohol. Electrode grid fixation was secured using circular adhesive bandages to ensure stable electrode–skin contact throughout the experimental protocol while minimizing movement artifacts. The reference electrodes were attached to the wrist using a water-moistened conductive strap.

For the FDS muscle, the elbow was slightly rotated medially with the hand palm facing upward. The reference lead line length (LLL) was determined by connecting the medial epicondyle of the humerus to the sulcus carpi at the wrist, and the central point of the electrode grid was positioned at 1/4 of the LLL from the medial epicondyle. Proper positioning was verified by asking the participant to flex their fingers against external resistance. For the EDC muscle, the upper arm was abducted laterally with the hand palm facing downward. The reference LLL was established by connecting the lateral epicondyle of the humerus to the midpoint between the styloid processes of the radius and ulna, and the central point of the electrode array was positioned at 1/4 of the LLL from the lateral epicondyle. Proper positioning was verified through finger extension.

Participants followed the instructions to replicate synchronously the dynamic, sinusoidal finger movements displayed by a virtual model of a bionic hand on a monitor placed in front of them (display size: 152.4 cm, positioned approximately 2 m in front of the participants; see Fig. 2), while seated comfortably in a chair with the elbow resting on a height-adjustable table. Tasks began with the fingers fully extended and comprised eight movement patterns: flexion and extension of the index finger, middle finger, and ring and pinky fingers simultaneously; thumb opposition and retraction; pinch grasp and release with the index finger and thumb, and with the middle finger and thumb; tripod pinch with the index, middle, and thumb fingers; and 5-finger grasp. Sinusoidal finger movements were displayed at 0.50 Hz and 0.75 Hz, with 16 trials per set. Each participant completed three sets (a total of 48 tasks). Each task lasted 45 s, with at least 30 s of rest between tasks. The order of the tasks was randomized, and HD sEMG signals were also recorded at rest to assess signal quality.

**FIGURE 2.**
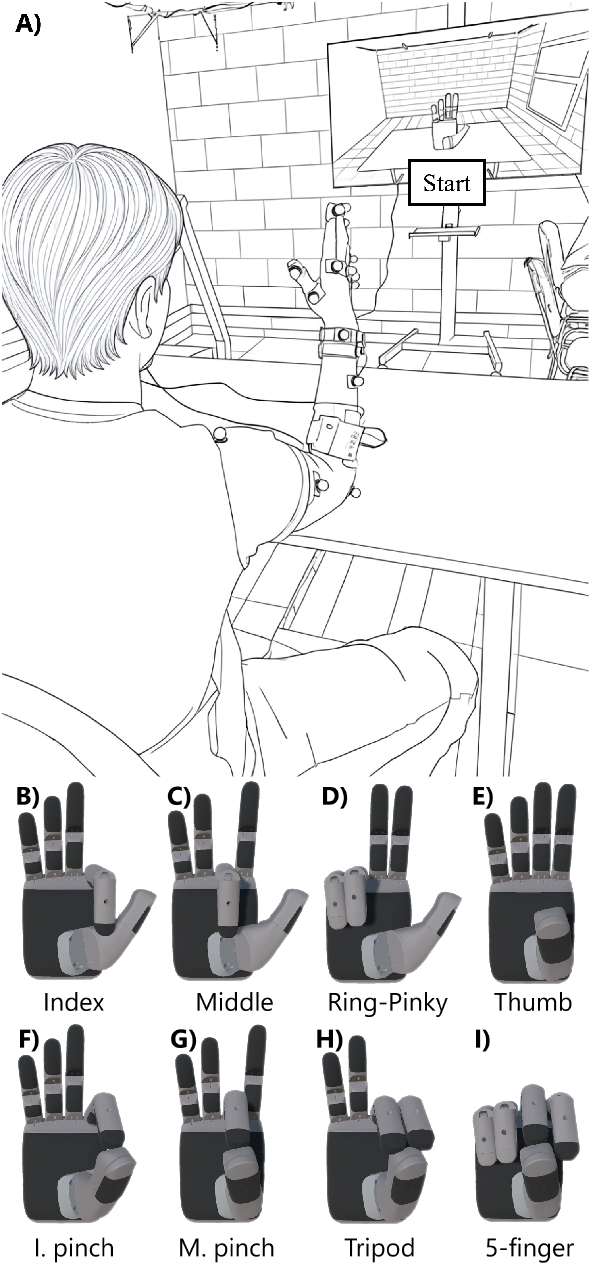
(A) Experimental setup. Each participant was instructed to follow sinusoidal dynamic movements displayed on a screen. HD sEMG of EDC and FDS muscles were recorded synchronously with the virtual bionic hand movements, along with hand markers to estimate joint angles by a Vicon motion capture system. Panels B to I illustrate the eight movements studied: (B) Index finger flexion; (C) Middle finger flexion; (D) Ring and pinky finger flexion; (E) Thumb opposition; F) Pinch grasp with the index finger; (G) Pinch grasp with the middle finger; (H) Tripod grasp; (I) 5-finger grasp.

A range-of-motion (ROM)-based calibration strategy was adopted to reduce inter-participant and inter-task variability in contraction force. Participants were instructed to perform each movement with a full ROM excursion required to accomplish the corresponding target movement. For non-grasping tasks, participants initiated each trial with the hand fully opened and then performed the required movement in a natural manner, without hyperextension or forceful flexion. The motion, therefore, extended from a standardized open-hand posture to the task-defined end posture (e.g., extension–flexion or opposition–reposition). For grasping tasks, finger flexion was performed until light fingertip contact was achieved, avoiding excessive force (as verified by the experimenter from the EMG interference pattern). In these tasks, participants were explicitly instructed to start with the thumb fully extended, as some tended to maintain the thumb in opposition while moving only the other digits. This procedure standardized movement execution and the associated muscle activation across participants, tasks, and sets.

The designed platform synchronized the start of HD sEMG signal recording, the virtual bionic hand movements displayed on the monitor, and the motion capture system (Fig. 3). Synchronization signals were transmitted over Wi-Fi using the TCP/IP protocol to the monitor interface, developed with Unity Engine (Unity Technologies, USA), and via the UDP/IP protocol to the Vicon Vero 2.2 motion capture system (Vicon Motion Systems, UK). Kinematic data were processed using a custom-made trigonometric model that estimates the 1-D carpometacarpal joint angle for the thumb and the metacar-pophalangeal joint angles for the remaining fingers [29].

**FIGURE 3.**
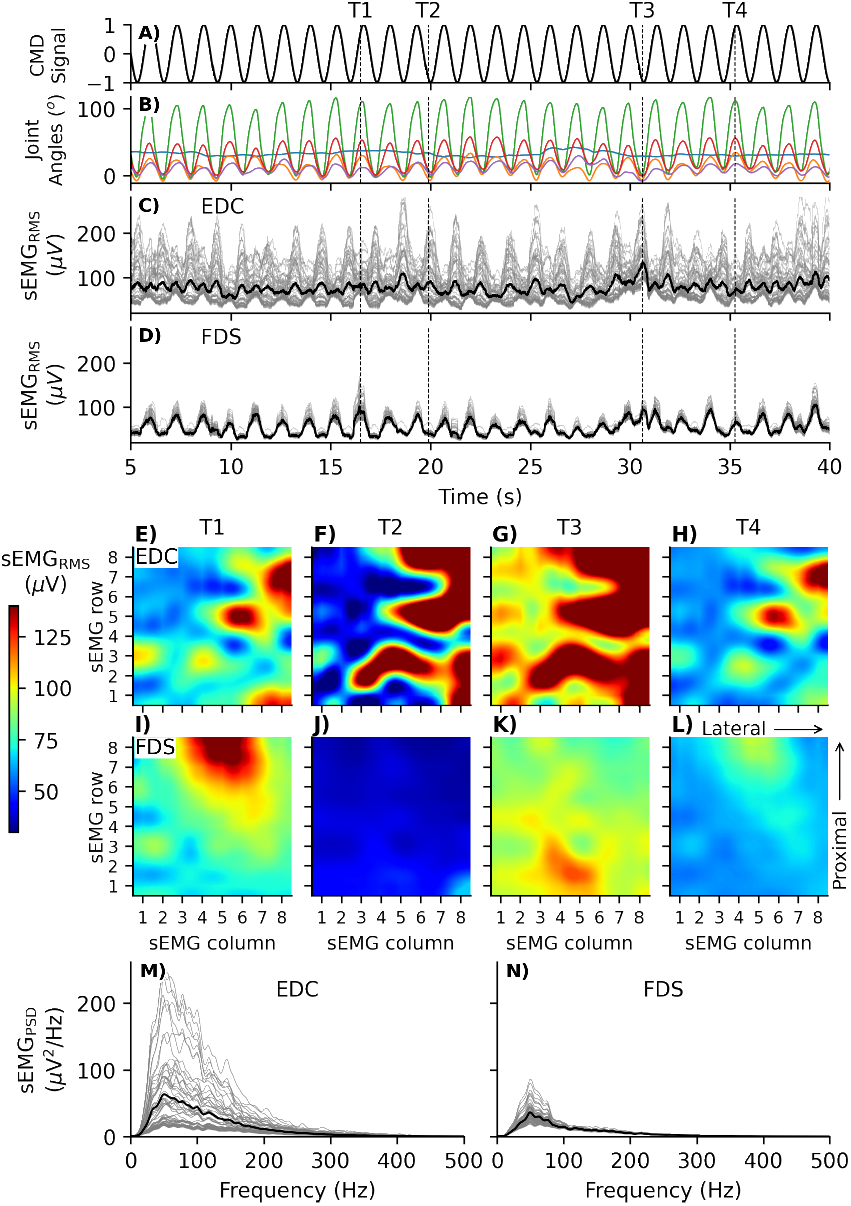
Example of the recordings obtained during the flexion-extension task of the middle finger at 0.75 Hz. (A) Command (CMD) signal controlling the position of the fingers of the virtual bionic hand displayed on the screen. (B) Metacarpophalangeal joint angles of the middle (green), ring (red), index (orange), thumb (blue), and pinky (purple) fingers. (C) sEMG_RMS_ from the electrode grid positioned over the EDC muscle (gray), along with the average sEMG_RMS_ across all channels (black). (D) sEMG_RMS_ from the electrode grid positioned over the FDS muscle (gray), and the corresponding average signal (black). (E) to (G) sEMG_RMS_ activation maps from the electrode grid over the EDC muscle. (I)-(L) sEMG_RMS_ activation maps from the electrode grid over the FDS muscle. (M) and (N) sEMG_PSD_ (Power Spectral Density) of the electrode grid over the EDC and FDS muscles, respectively (gray), along with their mean PSD (black).

### C. HD sEMG PROPERTIES

HD sEMG recordings with the new platform were analyzed in both the time and frequency domains. The signals underwent only the analog and digital filtering stages integrated into the system’s front-end, with no additional filtering applied during processing of the properties and quality metrics. The evaluation of HD sEMG characteristics included the maximum peak-to-peak amplitude of the sEMG (sEMG_P2P_), mean value of the sEMG_RMS_, coefficient of variation (CoV) of the sEMG_RMS_, and median frequency (MF) of the sEMG power spectral density (sEMG_PSD_). Calculation of the sEMG_RMS_ employed 300 ms windows. The PSD was estimated using Welch’s method with a 2-s Hanning window, without overlap.

### D. HD sEMG QUALITY ASSESSMENT

The assessment of HD sEMG data quality used metrics derived from sEMG_PSD_, which quantified the contamination level of each recording, including motion artifacts, power-line interference, noise, and intrinsic distortions of the acquisition system [30]. Six quality metrics were calculated (Tab. 2): Signal-to-Noise Ratio (SNR), Signal-to-Motion-artifact Ratio (SMR), Signal-to-Power-line-interference Ratio (SPR), Signal-to-High-frequency-noise Ratio (SHR), Spectrum Maximum-to-Minimum Drop in power density (SMMD), and Power Spectrum Deformation Ratio (PSDR). The evaluation of the results was based on previously defined acceptable levels reported in the literature, when available [31], [32]. The distribution of SNR values was analyzed based on the taxonomy proposed by [32], where values below 1.8 dB do not necessarily indicate unacceptable signals but a low SNR, suggesting a lack of relevant myoelectric activity for control applications; signals with SNR between 1.8 dB and 10 dB are acceptable; signals with SNR between 10 and 18 dB demonstrate minimized noise interference, rendering them highly suitable for precise control; and values above 18 dB indicate negligible noise, characterizing high-quality signals ideal for myoelectric control systems.

### E. PROPORTIONAL MOVEMENT INTENTION DECODING LOOP

The estimation time (*T*_*e*_) of the PMID loop and contribution of each stage (conditioning filters, RESA, RMS, NMF, Kalman, and CAN) were evaluated for analysis window lengths (*T*_*a*_) of 30, 60, 90, 120, and 150 ms. We calculated the accumulated time from the start of the acquisition window to the end of processing (*T*_*a*_ + *T*_*e*_) and the ratio *T*_*e*_*/T*_*a*_. This ratio indicates the degree of overlap between consecutive windows, as the step between them approximates *T*_*e*_. Values above 1 imply no overlap—risking loss of transient signal information, whereas values below 1 indicate overlapping windows, which enhance the temporal resolution.

**TABLE 2.** Spectral metrics used to evaluate the quality of myoelectric signals.

| Metric (Abbreviation) | Equation | Acceptable Level [31], [32] |
| --- | --- | --- |
| Signal-to-Noise Ratio (SNR) | $\frac{P_{\text{signal}}}{P_{\text{rest}}}$ | > 1.8 dB |
| Signal-to-Motion-artifact Ratio (SMR) | $\frac{P_{\text{signal}}}{P_{0-20 \text{ Hz}}}$ | > 12 dB |
| Signal-to-Power-line-interference Ratio (SPR) | $\frac{P_{\text{signal}}}{P_{60,120,\dots}}$ | N/A |
| Signal-to-High-frequency-noise Ratio (SHR) | $\frac{P_{\text{signal}}}{P_{500-1026 \text{ Hz}}}$ | > 15 dB |
| Spectrum Maximum-to-Minimum Drop in power density (SMMD) | $\frac{MPD_{\text{highest}}}{MPD_{\text{lowest}}}$ | > 30 dB |
| Power Spectrum Deformation Ratio (PSDR) | $\frac{(M_2/M_0)^{0.5}}{M_1/M_0}$ | < 1.4 |

$P_{\text{signal}}$ and $P_{\text{rest}}$ are the total signal power measured during the task and at rest, respectively. $P_{0-20 \text{ Hz}}$ represents a power below 20 Hz. $P_{60, 120, \dots \text{ Hz}}$ corresponds to the power at 60 Hz and its harmonics up to half of the sampling frequency. $P_{500-1026}$ denotes the sum of the power at frequencies greater than 500 Hz. $MPD$ represents the mean power density of 13 consecutive points shifted across all the frequencies. $M_n$ is the $n$ -th order spectral moment.

The CAN transmission time was measured 10 times using a RIGOL MSO5074 oscilloscope and an ESP32 WROOM-32E microcontroller. It was defined as the interval between the rising edge of a step signal generated by the MPSoC just before sending a CAN message and the rising edge of a signal generated by the microcontroller upon message reception.

For NMF training, the first 5 s of the HD sEMG recordings were discarded to avoid transients, and the following 20 s were used for model training. The analysis window (*T*_*a*_) was defined as the sliding window for the RMS calculation to ensure consistent temporal parameters during testing. The coefficient of determination (R^2^) was used to quantify the extent to which the NMF-reconstructed sEMG_RMS_ signals matched the original data.

During training, the RESA weight matrix was updated every 30 ms, and the final weights served as the initial values for testing. The last 20 s of each trial (testing interval) were used to assess the PMID loop’s ability to track oscillations in the command signal. During testing, the PMID loop processed consecutive windows of length *T*_*a*_ using the spatial component matrix learned during training (Fig. 4A–D).

**FIGURE 4.**
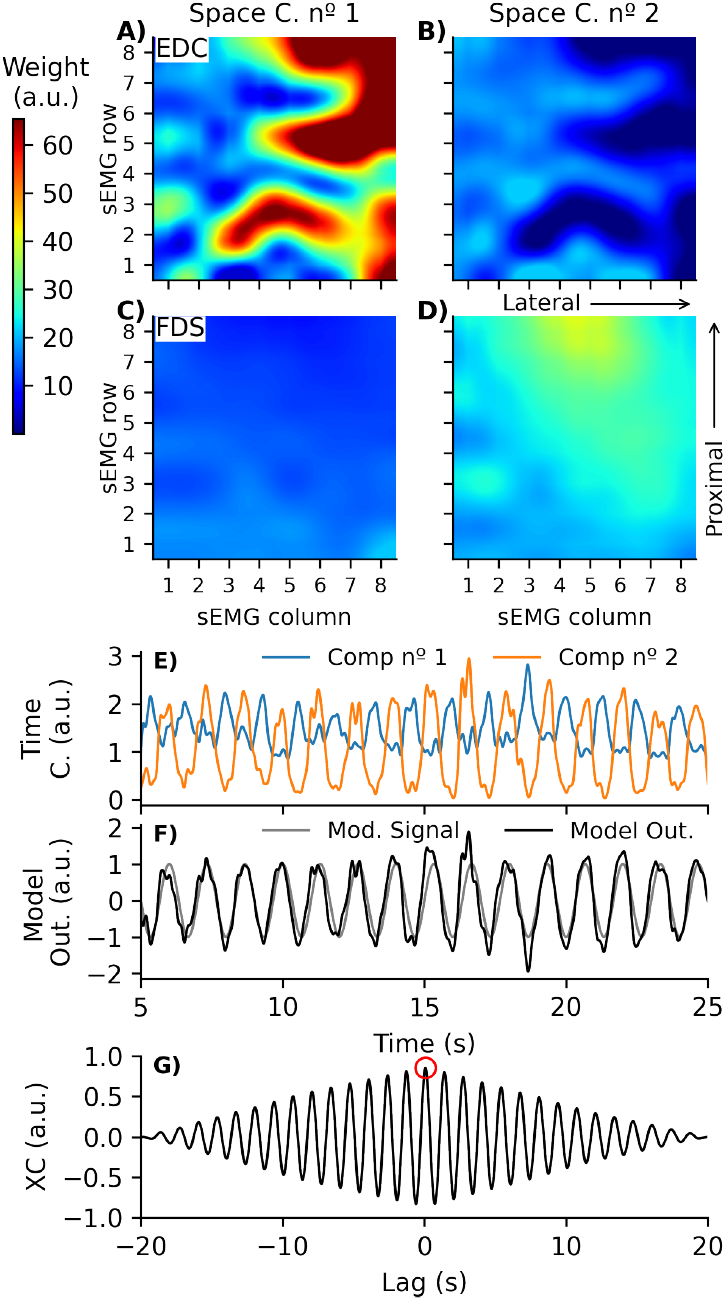
Example of the NMF decomposition and similarity evaluation between model output and command signals. (A and C) First spatial components extracted from the electrode grids over the EDC and FDS. (B and D) The second spatial components from the EDC and FDS muscles. (E) First and second temporal components. (F) Command signal of the virtual bionic hand (gray) and PMID loop output (black). (G) Normalized cross-correlation function between command and estimated signals (black) with XC_max_ highlighted (red circle).

### F. POWER CONSUMPTION AND BATTERY AUTONOMY ASSESSMENT

Battery autonomy was assessed through complete discharge tests conducted on three independent battery units. Each experiment consisted of continuous platform operation under full-load conditions, including simultaneous acquisition of 128-channel HD sEMG signals, real-time processing on the MPSoC (both the processing system and programmable logic), and graphical user interface updates, maintained until the battery reached the BMS-defined cutoff threshold. During discharge, the BMS recorded battery parameters at 30 s intervals via register interrogation. The monitored variables included total battery voltage, discharge current, temperature, and estimated time to empty (TtE). The average system power consumption was computed from the instantaneous power values obtained as the product of the measured voltage and current at each sampling point.

The accuracy of the BMS TtE estimation was assessed by computing, at each sampling time, the absolute error between the BMS-reported TtE and the actual remaining discharge time, calculated as the time interval from the current instant to the experimental endpoint. In addition, the *Report Power* tool from the Xilinx Vivado Design Suite (v2023.2) was used to estimate the power consumption of the MPSoC, providing separate power values for the processing system (PS) and programmable logic (PL). The remaining power consumption was attributed to the platform’s peripheral components.

### G. STATISTICAL ANALYSIS

Before statistical analysis, the normality of data from HD sEMG properties and the similarity between the estimated proportional movement intention and command signals were assessed using the D’Agostino-Pearson test. In both cases, the data failed the normality assumption across all metrics (*p* < 0.001), indicating the need to adopt nonparametric tests.

The influence of task type and task frequency on HD sEMG properties was analyzed using a Kruskal–Wallis test with multiple factors. Given the large sample size, all comparisons were statistically significant (*p* < 0.001) across all metrics. Therefore, the analysis focused on reporting the effect size using epsilon squared (*ε*^2^), which provides an effect size estimate that is independent of the sample size [33]. The effect size was considered negligible when *ε*^2^ < 1 %, small when 1 % ≤ *ε*^2^ < 8 %, medium when 8 % ≤ *ε*^2^ < 26 %, and large when *ε*^2^ ≥ 26 % [34].

The similarity between the estimated proportional movement intention and command signals was quantified by the maximum normalized cross-correlation (XC_max_) of the test data (Fig. 4G). The effect of *T*_*a*_ on XC_max_ was analyzed using the Kruskal–Wallis test. The effects of task type and frequency on XC_max_, training R^2^, and the lag at XC_max_ during testing were assessed using a multifactorial Kruskal–Wallis test at *T*_*a*_ = 60 ms, followed by Dunn’s post hoc test for pairwise comparisons.

All descriptive statistics are reported as the mean ± standard deviation. Statistical significance was set at *p* < 0.05.

## IV. RESULTS

### A. HD sEMG PROPERTIES

Of the 129,024 signals recorded during the platform evaluation (corresponding to all channels across participants, trials, and sets), only two channels (0.002 %) exceeded the amplifier’s linear range (± 5 mV). At rest, HD sEMG showed sEMG_P2P_ of 244.15 ± 253.14 *µ*V, sEMG_RMS_ of 15.62 ± 11.02 *µ*V, and sEMG_PSD_ MF of 128.90 ± 37.18 Hz. The low sEMG_RMS_ at resting confirmed negligible muscle activity and defined the baseline for SNR calculation.

Dynamic tasks at 0.75 Hz produced higher sEMG_P2P_ amplitudes (1.36 ± 0.64 mV) than at 0.50 Hz (1.13 ± 0.55 mV; Fig. 5A), with a small effect size of frequency (*ε*^2^ = 4.37 %). Mean sEMG_RMS_ followed the same trend: 82.60 ± 36.69 *µ*V at 0.75 Hz vs. 66.71 ± 29.07 *µ*V at 0.50 Hz (*ε*^2^ = 6.83 %). Task type had a medium effect size on sEMG_P2P_ (*ε*^2^ = 10.80 %) and a small effect size on mean sEMG_RMS_ (*ε*^2^ = 6.75 %). The interaction between task type and frequency had medium effect sizes (sEMG_P2P_: *ε*^2^ = 15.37 %; sEMG_RMS_: *ε*^2^ = 13.68 %). The 5-finger grasp task produced the highest mean sEMG_RMS_ (0.75 Hz: 96.34±47.25 *µ*V; 0.50 Hz: 73.29 ± 30.11 *µ*V), whereas the lowest values occurred in the Index (0.75 Hz: 68.65 ± 26.53 *µ*V) and Thumb (0.50 Hz: 55.93 ± 25.04 *µ*V) tasks (Fig. 5B).

**FIGURE 5.**
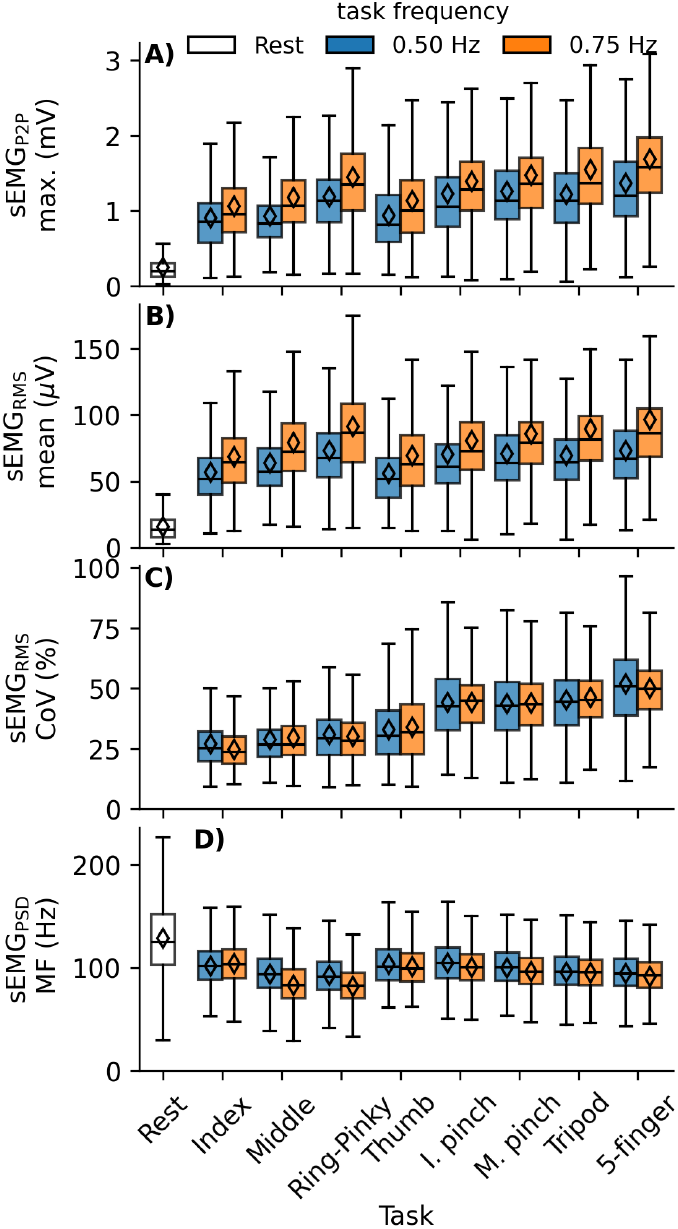
Distributions of HD sEMG signal properties by task type and movement frequency (*N* = 8, 064). (A) Peak-to-peak amplitude (sEMG_P2P_), (B) mean sEMG_RMS_, (C) coefficient of variation (CoV) of sEMG_RMS_, (D) median frequency (MF) of sEMG_PSD_.

Kruskal–Wallis analysis revealed a large effect of task type on sEMG_RMS_ CoV (*ε*^2^ = 35.28 %), a negligible effect of frequency (*ε*^2^ = 0.02 %), and a large task type and frequency interaction (*ε*^2^ = 35.47 %), indicating that CoV primarily depended on task type, with higher variability during grasping. The sEMG_RMS_ CoV was consistently high for the 5-finger grasp (0.50 Hz: 51.8 ± 18.0 %; 0.75 Hz: 50.1 ± 13.1 %) and low in single-finger tasks (e.g., Index at 0.75 Hz: 24.8 ± 7.7 %; Middle at 0.50 Hz: 28.7 ± 10.6 %).

The sEMG_PSD_ exhibited the expected monopolar pattern with power concentrated between 20–150 Hz [35], [36]. The Kruskal–Wallis test revealed small effect sizes for task type (*ε*^2^ = 7.38 %) and frequency (*ε*^2^ = 1.00 %), and a medium effect size for the interaction between task type and frequency (*ε*^2^ = 9.20 %). The sEMG_PSD_ MF decreased by approximately 10 Hz in Middle and Ring–Pinky tasks at 0.75 Hz vs. 0.50 Hz (e.g., Middle: 83.14 ± 22.84 Hz vs. 93.78 ± 21.58 Hz), whereas other tasks showed smaller changes (Index: 103.94 ± 20.75 Hz vs. 101.72 ± 22.98 Hz; Thumb: 100.80 ± 18.66 Hz vs. 103.72 ± 21.61 Hz; Fig. 5D).

### B. HD sEMG QUALITY ASSESSMENT

The analysis of the distribution of the quality metric values for the HD sEMG signals (Fig. 6) indicated that all metrics exceeded their respective acceptance thresholds. The SNR (Fig. 6A) showed a mean value of 13.93 ± 7.51 dB, reflecting the overall quality of the acquired signals. Only 2.76 ± 9.32 % of the channels exhibited an SNR < 1.8 dB, suggesting a small percentage of functionally irrelevant signals for control purposes. Approximately 30.58 ± 34.12 % of the signals fell within the 1.8 to 10 dB, indicating suitability for myoelectric control applications. The highest proportion of channels (39.66 ± 35.03 %) exhibited SNR values between 10 and 18 dB, a range where noise impact on extracted sEMG parameters is considered minimal. Finally, 27.00 ± 39.35 % of the channels showed an SNR above 18 dB, indicating negligible noise and high-quality signals suitable for myoelectric control. The SNR values close to the acceptance threshold were expected, given the low-intensity contractions during the dynamic movements employed in our study, with sEMG_RMS_ values below those observed in isometric tasks at 10 % of maximum voluntary contraction [37].

**FIGURE 6.**
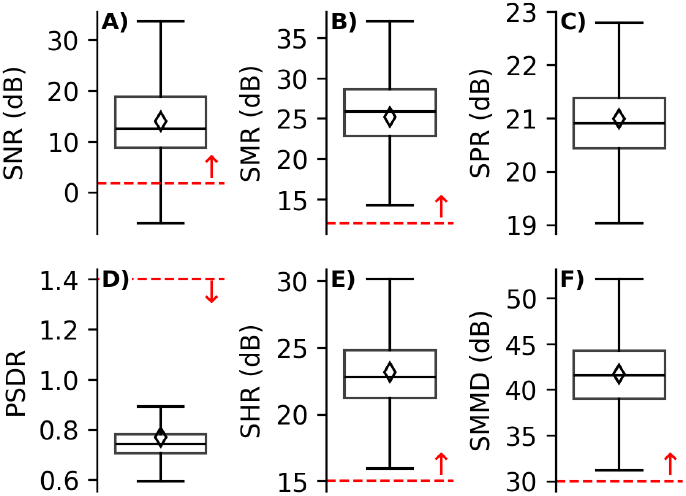
Distributions of spectral quality metrics of the HD sEMG signals. Red dashed lines and arrows are shown only when acceptable thresholds are reported in the literature [32]. Arrows indicate the direction of acceptable values. (A) Signal-to-Noise Ratio (SNR). (B) Signal-to-Motion Artifact Ratio (SMR). (C) Signal-to-Power Line Interference Ratio (SPR). (D) Power Spectrum Deformation Ratio (PSDR). (E) Signal-to-High Frequency Noise Ratio (SHR). (F) Spectrum Maximum-to-Minimum Drop in Power Density (SMMD).

The SMR metric (Fig. 6B) showed a value of 25.18 ± 5.18 dB, with only 2.44 ± 14.55 % of channels falling below the 12 dB threshold, suggesting that motion artifacts were minimal in the recorded data. The SPR values (Fig. 6C), which quantify power line interference at 60 Hz and its harmonics, were 20.99 ± 1.30 dB. While no established threshold exists for this metric in the literature, a visual examination of the sEMG_PSD_ reveals negligible interference. The PSDR metric (Fig. 6D) had a value of 0.77 ± 0.22, with 98.67 ± 11.03 % of channels below the recommended threshold of 1.4 [31], indicating a high spectral quality of the signals. The SHR metric (Fig. 6E) yielded a value of 23.16 ± 2.85 dB, with 99.81 ± 1.08 % of channels exceeding the 15 dB threshold, indicating a negligible presence of high-frequency noise in the signals. The SMMD metric achieved a mean value of 41.71 ± 4.69 dB, with 99.20 ± 1.86 % of channels surpassing the 30 dB threshold, indicating that nearly all signals exhibit power spectra properly sampled across the frequency range corresponding to sEMG signals.

### C. PROPORTIONAL MOVEMENT INTENTION DECODING LOOP

The developed platform successfully controlled the bionic hand in real time, with the PMID loop output effectively modulating the prosthetic hand actuators (Fig. 7; see supplementary videos). Fig. 8 summarizes the estimation time (*T*_*e*_) and total processing time (*T*_*a*_ + *T*_*e*_) of each PMID loop stage for different analysis windows (*T*_*a*_). Processing times for filters, RESA, and RMS increased with *T*_*a*_, while NMF, Kalman, and CAN remained nearly constant. RESA had the highest processing time (17.69 ± 0.82 ms to 35.34 ± 4.36 ms), followed by filtering (9.21 ± 0.17 ms to 22.34 ± 0.70 ms) and RMS (1.39 ± 0.04 ms to 6.26 ms). NMF, Kalman, and CAN times were 3.34 ± 0.31 ms, 0.05 ± 0.02 ms, and 1.79 ± 0.55 ms, respectively. *T*_*e*_ ranged from 33.30 ± 1.01 ms to 69.11 ± 4.47 ms. Total control latency (*T*_*a*_+*T*_*e*_) increased from 63.30 ± 1.01 ms (*T*_*a*_ = 30 ms) to 219.11 ± 4.47 ms (*T*_*a*_ = 150 ms). The *T*_*e*_*/T*_*a*_ ratio exceeded 1 only at *T*_*a*_ = 30 ms, indicating no overlap and potential loss of transient information. For *T*_*a*_ ≥ 60 ms, the ratio dropped below 1 (0.76–0.46), reflecting an increasing overlap and improved temporal resolution.

**FIGURE 7.**
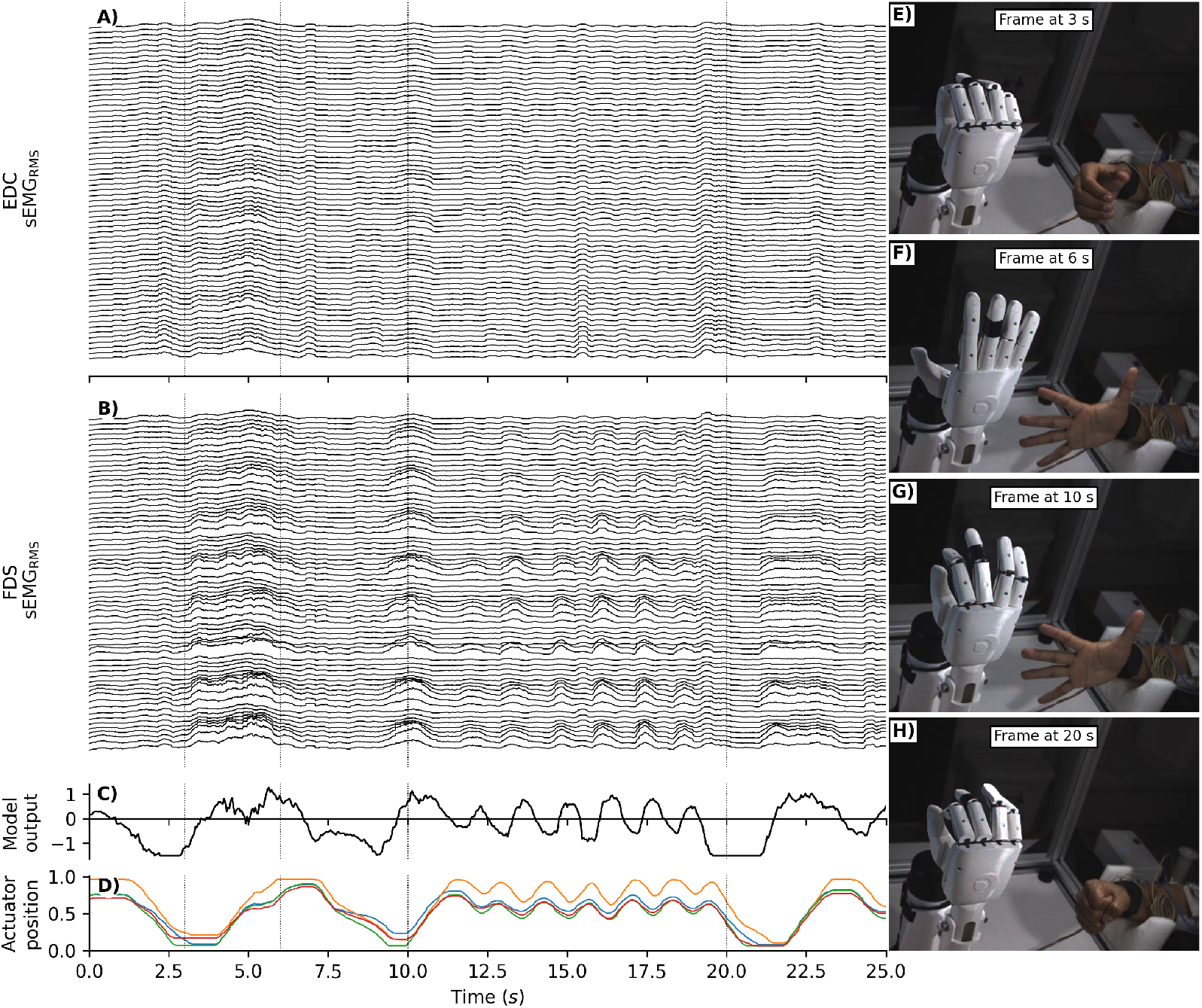
Illustrative example of real-time myoelectric control of the bionic hand during a 5-finger grasp task. (A) sEMG_RMS_ recorded from the electrode grid positioned over the *Extensor Digitorum Communis* (EDC) muscle. (B) sEMG_RMS_ from the electrode grid over the *Flexor Digitorum Superficialis* (FDS) muscle. (C) Output of the Proportional Movement Intention Decoding (PMID) loop. (D) Position of the bionic hand actuator shafts corresponding to the thumb (blue), index (orange), middle (green), and ring-pinky fingers (red); (E) to H) Video frames synchronized with the HD sEMG recordings, showing the bionic hand and the participant operating the myoelectric control system at 3 s, 6 s, 10 s, and 20 s, respectively (also indicated by vertical lines in the plots).

**FIGURE 8.**
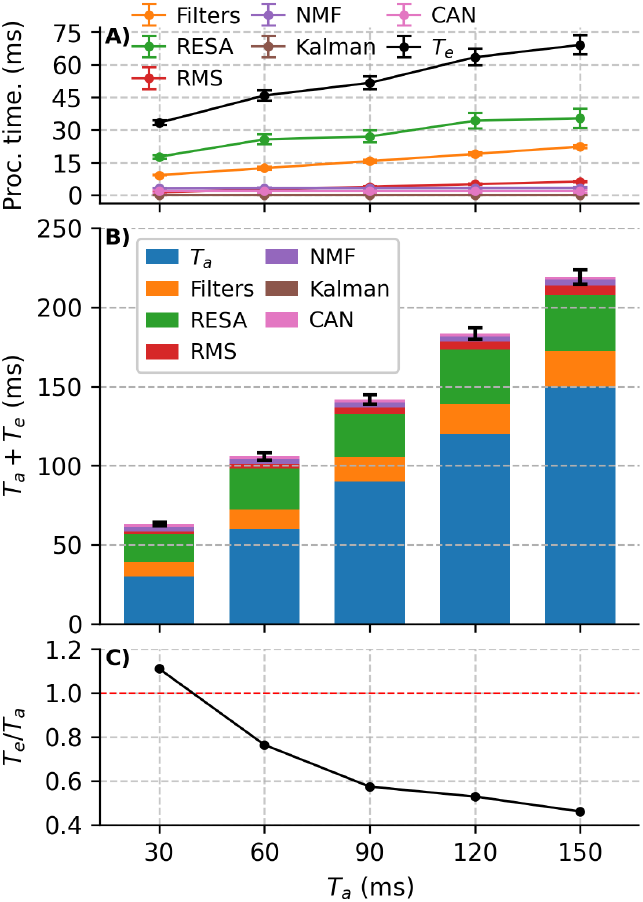
Processing performance of the Proportional Movement Intention Decoding (PMID) loop as a function of acquisition window (*T*_*a*_). (A) Processing time of each stage, with estimation time (*T*_*e*_) shown in black. (B) Total acquisition and estimation time (*T*_*a*_ + *T*_*e*_), with stacked bars representing the contribution of each processing stage. (C) Ratio of *T*_*e*_ to *T*_*a*_; the red dashed line indicates *T*_*e*_ = *T*_*a*_, with values below the line representing estimation times faster than acquisition. Error bars denote standard deviation. Sample size slightly decreased with *T*_*a*_ : *N* = 66,640 (30 ms), 66,560 (60 ms), 66,480 (90 ms), 66,400 (120 ms), and 66,320 (150 ms).

Fig. 9 shows R^2^ and XC_max_ distributions as a function of *T*_*a*_. R^2^ slightly increased with larger windows (0.89 ± 0.07 to 0.94 ± 0.07). XC_max_ during training rose from 0.51 ± 0.19 to 0.67 ± 0.19, likely due to reduced RMS noise with higher *T*_*a*_. XC_max_ for the test data remained unchanged (∼ 0.66 ± 0.20), with no significant differences across *T*_*a*_ (Kruskal–Wallis, *p* = 0.197, *ε*^2^ = 0.12 %), indicating that model generalization was unaffected.

**FIGURE 9.**
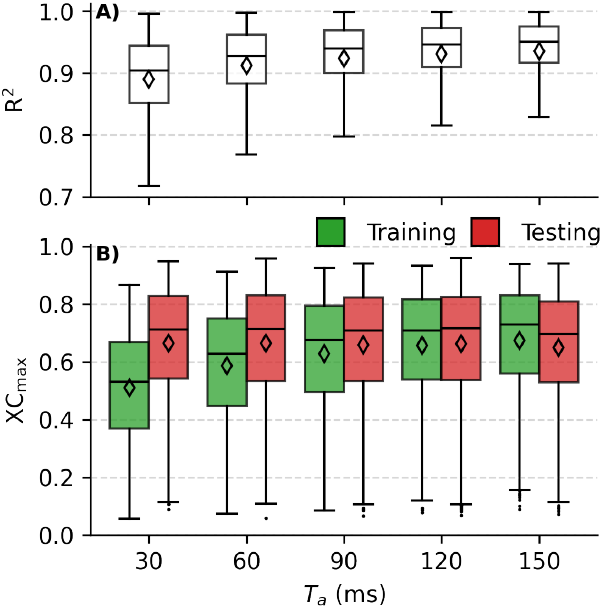
Performance metrics of the Proportional Movement Intention Decoding (PMID) loop as a function of acquisition windows (*T*_*a*_, *N* = 1, 008). (A) Distributions of the coefficient of determination (R^2^) for reconstructing HD sEMG signals from the NMF components. (B) Distributions of the maximum normalized cross-correlation values (XC_max_) between the estimated signals and the command signals for training (green) and testing (red) data. Diamonds denote mean values, boxes represent the interquartile range, and whiskers indicate minimum and maximum values (excluding outliers).

Fig. 10 presents the performance of the PMID loop across the motor tasks and the execution frequencies for *T*_*a*_ = 60 ms. The Kruskal–Wallis test revealed that task type had a large effect on R^2^ (*p* < 0.001, *ε*^2^ = 28.94 %), frequency had no significant effect (*p* = 0.053, *ε*^2^ = 0.37 %), and the interaction between task type and frequency was significant with a large effect size (*p* < 0.001, *ε*^2^ = 29.53 %), indicating that the influence of frequency depended on the specific task. Grasping tasks consistently showed the highest reconstruction quality (R^2^ = 0.95 ± 0.03 for the 5-finger grasp at 0.75 Hz). In contrast, non-grasping movements (Index, Middle, and Ring–Pinky tasks) exhibited lower performance (R^2^ = 0.87 ± 0.09 for index flexion–extension at 0.50 Hz).

Kruskal–Wallis analysis of XC_max_ for the test data revealed significant effects of task type (*p* < 0.001, *ε*^2^ = 11.70 %), frequency (*p* = 0.003, *ε*^2^ = 0.87 %), and the interaction between task type and frequency (*p* < 0.001, *ε*^2^ = 12.92 %). The Middle task at 0.50 Hz exhibited the highest similarity between the model output and command signal (0.80 ± 0.16). In contrast, the Index task at 0.75 Hz showed the lowest similarity (0.54 ± 0.22) compared with the other tasks.

**FIGURE 10.**
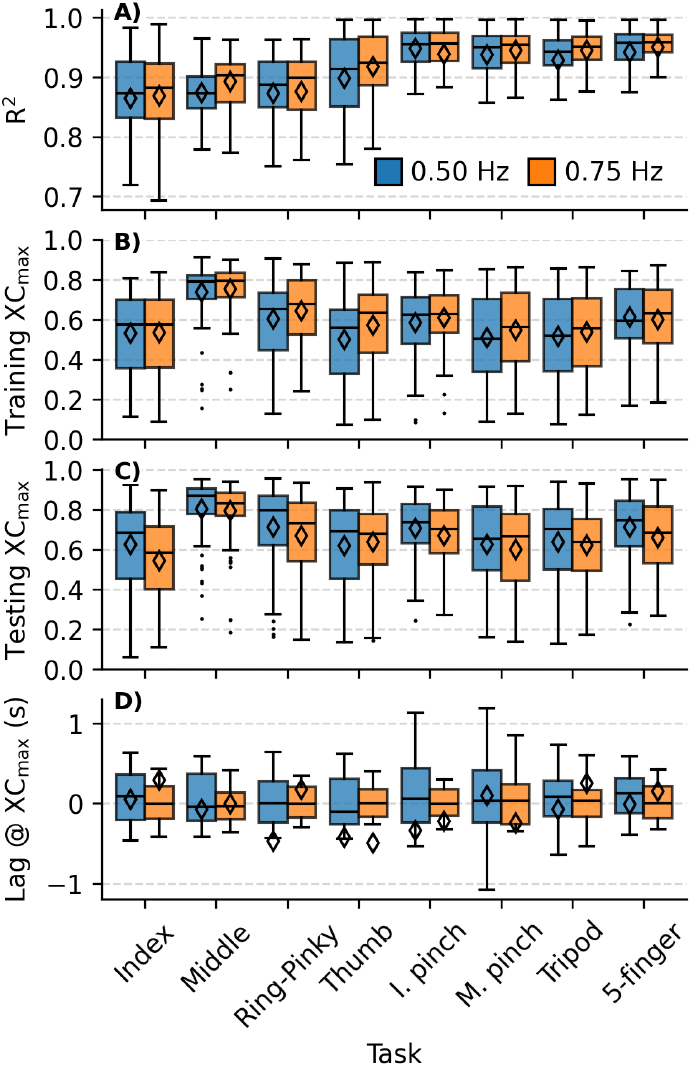
Distribution of performance metrics for the Proportional Movement Intention Decoding (PMID) loop across different motor tasks and execution frequencies, for *T*_*a*_ = 60 ms (*N* = 63). (A) Coefficient of determination (R^2^) for the reconstruction of HD sEMG signals from NMF components. (B) Maximum normalized cross-correlation value (XC_max_) for the training data. (C) XC_max_ for the test data. (D) Temporal lag at which XC_max_ occurs for the test data. Diamonds represent the mean values, boxes indicate the interquartile range, and whiskers show the minimum and maximum values (excluding outliers).

Analysis of the cross-correlation lags showed values near zero for all tasks and frequencies (0.02 ± 2.09 s). Kruskal–Wallis test indicated no significant effects of task type (*p* = 0.578, *ε*^2^ = 0.56 %), frequency (*p* = 0.459, *ε*^2^ = 0.05 %), or their interaction (*p* = 0.754, *ε*^2^ = 1.09 %), confirming the minimal temporal offset in the model output.

### D. POWER CONSUMPTION AND BATTERY AUTONOMY

Battery temperature remained within a narrow operating range throughout all experiments, exhibiting a mean value of 27.80 ± 0.91 °C (range: 26.25 °C to 30.45 °C), thereby confirming safe thermal operation. The BMS measured power during discharge at a mean value of 6.02 ± 0.11 W, with power distribution analysis via Vivado *Report Power* revealing that the MPSoC consumed 3.30 W (approximately 55 % of total system consumption). Of this, the PS consumed 2.95 W (89 %) and the PL consumed 0.35 W (11 %), while peripheral platform components accounted for the remaining 2.71 W. The time to empty (TtE) error analysis revealed a mean value of 1.03 ± 0.38 h across complete discharge cycles, indicating that the BMS consistently predicted longer operational time than the system actually achieved until complete discharge. Across all three discharge experiments, the system sustained continuous operation until BMS cutoff, yielding a mean battery autonomy of 11 h and 45 min.

## V. DISCUSSION

### A. RESEARCH PLATFORM FOR EDGE-BASED REAL-TIME MYOELECTRIC CONTROL

The primary contribution of this work is the development and validation of a wearable platform designed to support real-time myoelectric control and the research and development of new control algorithms using HD sEMG. By adopting a heterogeneous computing architecture that integrates the analog front-end for HD sEMG acquisition with an adaptive MPSoC, the proposed system performs signal acquisition and processing entirely on-device, eliminating the dependence on external computation resources commonly reported in the literature [38], [39], which inherently reduces privacy risks by avoiding off-device data transmission and external data handling. This design enables continuous multichannel signal capture and low-latency real-time processing under realistic usage conditions, enhancing system autonomy and directly addressing key challenges in wearable experimentation, such as connectivity constraints, mobility limitations, and reliance on external computing infrastructure. Consequently, the platform is well-suited for edge-based myoelectric research [40], [41].

In line with this objective, rather than optimizing a single performance metric, the proposed platform is designed to balance channel density, processing capability, autonomy, and actuation support within a wearable experimental system. This system-level perspective provides a unified environment for implementing and evaluating complete, computationally demanding myoelectric control pipelines directly on the device, including advanced signal conditioning, high-dimensional feature extraction, learning-based algorithms, and closed-loop control strategies. By supporting these pipelines under realistic real-time and resource constraints, the platform enables the rigorous assessment of next-generation myoelectric control methods. The heterogeneous MPSoC architecture is a key enabler of the proposed platform, providing the computational flexibility required to support a wide range of signal processing and control strategies within a single wearable system. By prioritizing reconfiguration and algorithmic diversity over task-specific optimization, the platform enables systematic development and benchmarking of myoelectric control approaches. This design choice makes the system particularly well-suited for exploratory research and comparative studies, while explicitly accepting higher power consumption relative to application-specific implementations.

Latency characterization further reinforces the platform’s suitability for real-time myoelectric control. By explicitly measuring the end-to-end latency of the complete acquisition and processing chain, the system provides a realistic assessment of its performance in closed-loop control scenarios. This is critical for evaluating control performance, responsiveness, and user perception, which are fundamental aspects of practical myoelectric interfaces [42]. Also, the achieved battery autonomy of 11.75 h is particularly relevant for experimental protocols that require extended and uninterrupted operation. This operating time supports long acquisition sessions and realistic user studies, making the platform suitable for both laboratory experimentation and real-world evaluations of myoelectric control strategies.

Overall, the experimental results support the three main architectural design choices of the proposed platform. First, the deterministic FPGA-based acquisition successfully handled 128-channel HD sEMG at 2052.52 Hz/channel with zero data loss across all 129,024 recorded signals, demonstrating that the throughput bottleneck commonly observed in MCU-based systems has been overcome. Second, the embedded processing achieved end-to-end latencies of 63.3–219.1 ms with predictable timing, confirming the platform’s ability to support real-time signal processing and control pipelines. Third, the heterogeneous architecture’s measured power consumption (6.02 W total, with 3.30 W for the MPSoC) and 11.75-hour battery autonomy demonstrate sufficient computational headroom. Notably, the quad-core A53 APU, Mali-400 GPU, and dual R5F cores provide the scalability required to host future deep learning algorithms while maintaining wearable form factor constraints.

### B. COMPARISON WITH STATE-OF-THE-ART HD sEMG PLATFORMS

To systematically compare the features and performance of the proposed platform, we present a benchmark analysis against recently developed HD sEMG systems reported in the literature (Tab. 3). The comparison is organized along three dimensions: system-level specifications, analog front-end characteristics, and digital processing and communication capabilities. The analysis highlights three core advantages of the proposed architecture: (1) Throughput scalability: the dual RHD2164 configuration with an FPGA-based LVDS DDR SPI interface sustains 128-channel acquisition without CPU intervention, addressing the channel scalability limitations commonly associated with MCU-centered approaches; (2) Edge autonomy: unlike systems requiring external PC processing [38], [39] or hybrid SoC+PC architectures [13], the MPSoC enables fully on-device acquisition, processing, and actuation control, with experimentally measured end-to-end latency; (3) Algorithmic extensibility: the heterogeneous computing fabric (quad A53, Mali-400 GPU, dual R5F, and FPGA) offers the computational diversity and memory band-width needed to support future integration of convolutional neural networks, recurrent architectures, and real-time deep learning models directly on the wearable device.

Importantly, while application-specific implementations such as Moin et al. [43] achieve substantially lower power consumption (148 mW) through fixed-function ASIC designs, the proposed MPSoC-based architecture intentionally emphasizes computational headroom and algorithmic flexibility over aggressive power optimization. The PYNQ/Linux framework facilitates rapid prototyping and evaluation of a wide range of decoding strategies—from classical signal processing to deep learning—without requiring hardware modification. In this sense, the platform is better interpreted as a flexible research testbed rather than a task-optimized commercial solution.

### C. HD sEMG PROPERTIES AND QUALITY ASSESSMENT

The acquired signals exhibited amplitudes well below the full scale of the acquisition system, indicating an operational margin well within the linear range, with no evidence of saturation for the analyzed tasks. However, more intense muscle contractions, approaching maximum voluntary contraction, were not assessed here. Despite relatively low average sEMG_RMS_ values, consistent variations across tasks were observed, as reflected in the coefficient of variation (sEMG_RMS_ CoV), demonstrating the system’s sensitivity to varying levels of muscle activation across motor tasks. The reduced amplitude observed during thumb opposition movements likely arises from the predominant involvement of the intrinsic hand muscles, which are located distally within the hand and originate near the carpal bones. Because these muscles lie far from the electrode grids placed on the proximal forearm, the sEMG signals they generate are attenuated compared with signals from the more proximal and thicker extrinsic forearm muscles [45].

Higher mean sEMG_RMS_ values at increased task frequencies reflect increased movement cadence, which requires greater muscle recruitment to execute faster movements. This increased activation results in larger signal amplitudes, consistent with evidence showing a strong correlation between movement speed and surface EMG activity, where higher velocities systematically elicit stronger muscular responses [46].

Overall, the spectral quality metrics of the signals were quite satisfactory, showing negligible contamination by noise and artifacts, surpassing the minimum (or maximum for the PSDR) acceptability thresholds suggested in the literature [32]. Therefore, this initial investigation confirmed that the developed platform is well-suited for real-time myoelectric control applications and neurophysiological analysis of HD sEMG signals.

### D. PROPORTIONAL MOVEMENT INTENTION DECODING LOOP

The PMID loop demonstrated that the system successfully reconstructed the dynamics of the modulating signals, achieving high R^2^ and XC_max_ values, particularly in grasping tasks. However, these findings do not necessarily imply higher performance in motor tasks representing daily activities [17]. The higher R^2^ values observed in grasping tasks likely resulted from their reduced synergistic control, which facilitated the separability of 1-DoF patterns during the NMF stage. In contrast, non-grasping movements often require additional muscular contractions to stabilize fingers that are not actively engaged in the task [47]. These postural contractions distribute control more broadly and induce multimodal motor unit activity shaped by the hand configuration. Beyond physiological complexity, the observed performance degradation is also fundamentally constrained by the use of NMF as a linear decomposition method, which cannot fully capture the inherently non-linear relationships present in biological neuromuscular signals. Unlike the low-dimensional synergistic control characteristic of grasping, this may account for the lower R^2^ values observed when modeling single-finger movements [48], [49].

For the tasks analyzed, which involved cyclostationary signals, the cross-correlation function exhibited multiple peaks separated by integer multiples of the signal period. Consequently, XC_max_ values occurring at lags exceeding the command signal period may result from the signal’s inherent periodicity or intrinsic noise, rather than representing a true physiological delay. This interpretation aligns with the theoretical foundations outlined in the literature, which highlights the phase ambiguity inherent in periodic signals and the resulting limitations in accurately estimating time delays [50], [51].

The discrepancy between the lower explained variance (R^2^) in the reconstruction of sEMG_RMS_ signals and the strong similarity (XC_max_ for the test data) between the model output and command signal observed in the Middle task reflects an intrinsic limitation of this measure. Surface EMG amplitude metrics such as sEMG_RMS_ are more directly related to the amplitude of motor unit action potentials than to the underlying motor unit activity modulation, thus providing only a coarse approximation of the neural drive for muscle activation [52]. These limitations underscore the necessity for high-performance hardware capable of supporting more complex non-linear models, such as advanced decomposition algorithms and deep learning approaches, which can better represent the intricate dynamics of individual finger control.

**TABLE 3.** Comparative analysis of recent high-density surface electromyography (HD sEMG) acquisition platforms, highlighting system-level, analog front-end, and digital signal processing characteristics.

| Feature | Cerone et al. (2019) [38] | Moin et al. (2021) [43] | Mannatunga et al. (2020) [13] | Koutsoftidis et al. (2022) [39] | This work |
| --- | --- | --- | --- | --- | --- |
| *System Specifications* |  |  |  |  |  |
| Primary application | sEMG monitoring | Gesture recognition | Multimodal biosignal acquisition | Motor unit decomposition | Real-time myoelectric control |
| System architecture | Modular (32-ch) | Monolithic (64-ch) | Monolithic (128-ch) | Modular (4×32-ch) | Modular (2×64-ch + PU) |
| Portability | Body-worn module | Fully body-worn | Benchtop system | Portable, external PU | Fully body-worn |
| Dimensions (mm)* | 34×30×15 | — | AFE: 76.5×69<br>PU: 160×135 | 100×80 (per AFE) | AFE: 61×32×9.5<br>PU: 170×100×75 |
| Total weight (g) | 16.7 | 26 | — | — | AFE: 19<br>PU+Battery: 961 |
| Reported power (mW) | 393–440 | 148 | 2850 | <2000 | — |
| Full-load power (mW) | — | — | — | — | 6020 |
| Battery autonomy (h) | 5.0 | 6.0 | — | >4.0 | 11.75 |
| *Analog Front-End* |  |  |  |  |  |
| Amplifier | RHD2132 | Custom IC [44] | RHA2132 | AD8295 + ADS1298 | RHD2164 |
| Number of channels | 32 | 64 | 128 | 32–128 | 64–128 |
| Total analog gain (V/V) | 192 | — | 200 | 120.2 | 200 |
| Analog filtering | 0.1–20 kHz<sup>a</sup> | DC-coupled | 0.1–20 kHz<sup>a</sup> | 23 Hz–4 kHz | 0.1–20 kHz<sup>a</sup> |
| ADC resolution (bits) | 16 | 15 | 18 | 24 | 16 |
| Sampling rate (kHz/ch) | 2.048 (max: 30.0) | 1.0 | 15.0 (max: 31.25) | 2.0 (max: 8.0) | 2.053 (max: 30.0) |
| *Digital Signal Processing and Communication* |  |  |  |  |  |
| Processing paradigm | External (PC/mobile) | Edge | Hybrid (SoC+PC) | External (PC) | Edge (MPSoC) |
| Processor | External host (PC/mobile) | ARM Cortex-M3 + FPGA | Dual ARM Cortex-A9 + FPGA | External host (PC) | Quad A53 + FPGA + Mali-400 + Dual R5F |
| AFE interface | SPI (20 MHz, DMA) | Custom digital bus | FMC connector | Daisy-chain SPI | LVDS DDR SPI (DMA) |
| Connectivity and host / actuation interfaces | Wi-Fi (TCP/IP) | BLE (2.4 GHz, nRF51822) | Ethernet (1 Gbps) | USB 2.0 (isolated) | Wi-Fi (TCP/IP, UDP/IP), CAN |
| Partial latency (ms) ** | 12 ± 2<sup>b</sup> | 250<sup>c</sup> | 1.07<sup>d</sup> | < 35<sup>e</sup> | — |
| End-to-end latency (ms) | — | — | — | — | 63.3–219.1<sup>f</sup> |

\* Dimensions are given as reported in the original works, either as $L \times W$ or $L \times W \times H$ .
\*\* Latency values follow the definitions used in the original publications; most report partial latency components rather than full end-to-end latency. The proposed platform reports total end-to-end latency.
<sup>a</sup> Typical bandpass configuration: 10–500 Hz for sEMG applications.
<sup>b</sup> Measurement focused on the Wi-Fi link latency between the sensor and the host for real-time visualization.
<sup>c</sup> Value determined by the 250 ms sliding window required for gesture inference.
<sup>d</sup> Main contribution due to the FIFO buffer filling time for Ethernet transmission.
<sup>e</sup> Communication latency through the isolated USB 2.0 interface for PC-based processing.
<sup>f</sup> End-to-end latency dependent on the selected processing window length ( $T_a$ ) (30–150 ms).
— Not reported in the original publication or not applicable.
Abbreviations: AFE, analog front-end; PU, processing unit; ch, channels; IC, integrated circuit; DC, direct current; ADC, analog-to-digital converter; kHz/ch, kilohertz per channel; FPGA, field-programmable gate array; SoC, system-on-chip; MPSoC, multiprocessor system-on-chip; GPU, graphics processing unit; DMA, direct memory access; SPI, serial peripheral interface; LVDS, low-voltage differential signaling; DDR, double data rate; TCP, Transmission Control Protocol; IP, Internet Protocol; BLE, Bluetooth Low Energy; USB, Universal Serial Bus; UDP, User Datagram Protocol; CAN, Controller Area Network; PC, personal computer; ARM, Advanced RISC Machines; FIFO, first-in first-out; FMC, FPGA Mezzanine Card; Gbps, gigabits per second.

### E. FUTURE WORK

Future work may explore two main research directions enabled by the proposed platform. First, the design and implementation of optimized algorithms for simultaneous and proportional decoding of 4-DoF finger movements, leveraging the combined use of FPGA and GPU resources. Such algorithms could target semi-supervised learning paradigms tailored to clinical populations, particularly in scenarios where kinematic ground-truth data are limited, to improve decoding performance and usability in real-world rehabilitation contexts. Second, the integration of the analog front-end and bionic-hand components into individualized prosthetic socket systems for transradial amputee users. Patient-specific socket designs could ensure appropriate fit, comfort, and electrode placement on the residual forearm, enabling functional evaluation of the complete system under conditions that closely resemble daily-life activities and clinical use.

Additional future research opportunities may explore the full capabilities of the RHD2164 front-end, which natively supports impedance analysis and provides auxiliary analog inputs that facilitate the integration of additional sensors such as inertial measurement units (IMUs). In particular, the RHD2164’s capability to perform impedance measurements across multiple frequencies and channels enables the development of methods to mitigate signal non-stationarities caused by variations in skin–electrode impedance. Such approaches could support continuous monitoring of electrode contact quality and adaptive compensation strategies to improve signal reliability. Furthermore, the integration of IMUs to mitigate arm position effects, recognized as a major challenge for reliable hand gesture recognition in practical myoelectric control systems [53], may further enhance decoding robustness. The fusion of HD sEMG and inertial data has the potential to improve stability across different arm postures and dynamic conditions, thereby increasing the overall reliability of the proposed wearable platform.

## VI. CONCLUSION

This work presents the design and validation of a wearable HD sEMG acquisition and processing platform based on a heterogeneous architecture, enabling real-time estimation of proportional movement intention with 1-DoF. The system consistently produced outputs that closely matched the reference command signals during specific finger motor tasks, while maintaining processing latencies well within real-time control requirements. Although the current embedded implementation is limited to a single-DoF control and does not yet support coordinated and proportional multi-finger movements, the proposed system was conceived to be scalable and computationally flexible. Its heterogeneous MPSoC-based architecture and advanced analog front-end capabilities provide a solid technological basis for the incorporation of more sophisticated decoding strategies, multimodal sensing, and clinically oriented control paradigms. Overall, the results indicate that the proposed platform constitutes an extensible foundation for the next generation of wearable myoelectric systems by overcoming throughput bottlenecks through FPGA-based acquisition of 128-channel data, enabling standalone edge computing that eliminates communication latency and privacy risks, and providing the scalability required to host future deep learning algorithms.

## Supporting information

Supplemental Video 1 - 5-finger control

Supplemental Video 2 - 5-finger control

Supplemental Video 3 - Index control

Supplemental Video 4 - Index control

## Data Availability

All data produced in the present study are available upon reasonable request to the authors

## VII. ACKNOWLEDGMENT

The authors would like to thank Mr. Flavio Renato Santos, Mr. Nelio Hilton Gava, and Mr. Glaudson Frade Assunção from the Main Design Lab, Center for Biomedical Engineering, UNICAMP, for their technical support in designing and fabricating the platform and the bionic hand prototypes.

**RICARDO G. MOLINARI** received the Ph.D. degree in Electrical Engineering from the University of Campinas (UNICAMP), Brazil, in 2025. He is currently a Postdoctoral Researcher at the Center for Biomedical Engineering, UNICAMP. He also holds an M.Sc. in Electrical Engineering (2021) and a B.Eng. in Control and Automation Engineering (2020) from UNICAMP, and previously earned a B.Eng. in Civil Engineering from São Paulo State University (UNESP, 2010). His research focuses on computational neuroscience, biological signal processing, and the development of assistive technologies for motor rehabilitation. His work integrates high-density surface electromyography, multiscale neuromuscular modeling, and real-time embedded systems for proportional and simultaneous control of upper-limb prostheses.

**VALERIA AVILÉS-CARRILLO** received the B.S. degree in Mechatronics Engineering from the Universidad Militar Nueva Granada (UMNG), Bogotá, Colombia, in 2021, and an M.S. degree in Mechanical Engineering from the University of Campinas (UNICAMP), Brazil, in 2023. She is currently pursuing a Ph.D. degree in Electrical Engineering at UNICAMP under the supervision of Prof. Leonardo Abdala Elias. From 2021 to 2022, she worked as a Research Assistant at UMNG, where she developed simulation and implementation of advanced control strategies for a hexacopter-type unmanned aerial vehicle. Her research interests include mechatronics, robotics, control systems, and biomedical engineering, with a focus on human–machine interfaces.

**GUILHERME A. G. DE VILLA** received the B.S. degree in Electrical Engineering from the Federal University of Goiás, Brazil, in 2016, and the M.S. and Ph.D. degrees in Electrical and Computer Engineering from the same institution in 2019 and 2023, respectively. He completed postdoctoral training in Biomedical Engineering at the University of Campinas (UNICAMP), Brazil, in 2025. Since 2025, he has been an Assistant Professor at the State University of Goiás. Dr. De Villa has authored several articles and book chapters in biomechanics and biomedical engineering. His research interests include neuromechanical modeling of human movement, biomechanical signal processing, coordination variability analysis, nonlinear motor control dynamics, and neuroengineering approaches for orthotic and prosthetic device development.

**LEONARDO A. ELIAS** (M’24-SM’25) received the Ph.D. degree in Biomedical Engineering from the University of São Paulo, Brazil, in 2013. He was a Postdoctoral Fellow at the Biomedical Engineering Laboratory, University of São Paulo (2013-2014), and a Visiting Researcher at the Department of Neurorehabilitation Engineering, Georg-August University (Goettingen, Germany, 2015). He is currently an Associate Professor at the Department of Electronics and Biomedical Engineering, School of Electrical and Computer Engineering, University of Campinas. He is the Founder Director of the Neural Engineering Research Laboratory at the Center for Biomedical Engineering, University of Campinas. His research in neural engineering focuses on theoretical and computational, as well as experimental, investigations that aim to understand how different elements of the neuromusculoskeletal system interact during normal and pathological control of human movement. Additionally, based on basic research, his group designs new technologies for motor neurorehabilitation of patients with neurological disorders and amputees. He is a Research Productivity Fellow of CNPq (Brazilian National Science Foundation). He is a member of the International MotoNeuron Society, the IEEE Engineering in Medicine and Biology Society, the Brazilian Society for the Progress of Science, and the Brazilian Society of Biomedical Engineering. He is a member of the Board of Reviewing Editors of the eLife journal.

